# Under detection of depression in primary care settings in low and middle-income countries: A systematic review and meta-analysis

**DOI:** 10.1101/2020.03.20.20039628

**Authors:** Abebaw Fekadu, Mekdes Demissie, Rahel Berhane, Girmay Medhin, Teserra Bitew, Maji Hailemariam, Abebaw Minaye, Kassahun Habtamu, Barkot Milkias, Inge Petersen, Vikram Patel, Anthony J Cleare, Rosie Mayston, Graham Thornicroft, Atalay Alem, Charlotte Hanlon, Martin Prince

**Author notes:** **Authors’ email address** Abebaw Fekadu. **Authors’ email address:** Mekdes Demissie, Rahel Berhane, Girmay Medhin, Tesera Bitew, Maji Hailemariam, Abebaw Minaye, Kasahun Habtamu, Barkot Milkias, Inge Petersen, Vikram Patel, Anthony J Cleare, Rosie Mayston, Graham Thornicroft, Atalay Alem, Charlotte Hanlon, Martin Prince.

## Abstract

**Objective:** Depression is the commonest mental disorder in primary care but is poor identified. The objective of this review was to determine the level of detection of depression by primary care clinicians and its determinants in studies from low-and middle-income countries (LMICs).

**Methods:** 

**Design:** Systematic review and meta-analysis. Review protocol was registered in the PROSPERO database (CRD42016039704).

**Databases:** PubMed, PsycINFO, Medline, EMBASE, LILAC and AJOL.

**Quality assessment:** Risk of bias within studies evaluated with the Effective Public Health Practice Project (EPHPP).

**Synthesis:** “Gold standard” diagnosis for the purposes of this review were based on the 9-item Patient Health Questionnaire (PHQ-9; cutoff scores of 5 and 10), structured interview or expert diagnosis. Meta-analysis was conducted excluding studies on special populations. Analysis of pooled data were stratified by diagnostic approaches.

**Results:** A total of 2223 non-duplicate publications were screened. Ten publications, from two multi-country studies and eight single country studies, making 18 country level reports, were included. One of the multi-country studies used an enriched sample of screen positive participants. Overall methodological quality of the studies was good. Depression detection was 0.0% in five reports and <12% in another five. The pooled detection for two reports that used PHQ-9 at a cutoff point of 5 (combined sample size = 1426) was 3.9% (95% CI = 2.3%, 5.5%); in the four reports that used PHQ-9 cutoff score of 10 (combined sample size =5481), the pooled detection was 7.0% (95% CI = 3.9%, 10.2%). For the enriched sample, the pooled detection was 43.5 % (95% CI: 25.7%, 61.0%). Severity of depression and suicidality were significantly associated with detection.

**Conclusions:** The extremely low detection of depression by primary care clinicians poses a serious threat to scaling up mental healthcare in LMICs. Interventions to improve detection should be prioritized.

**Strength and limitation of study:**

▸ This is the first review of detection of depression in LMIC settings
▸ The review was comprehensive in terms of databases searched
▸ Screening tools were used as gold standards, which may lead to overestimation of prevalence and underestimation of detection
▸ The small number of studies and the use of different instruments and cutoff
▸ points precluded exploration of sources of heterogeneity
▸ The review does not include studies on distress or sub-threshold depression

## BACKGROUND

Depression is a major public health problem associated with impaired quality of life, disability and substantial healthcare costs^1^. It is a relatively common condition in primary health care (PHC), affecting up to 20% of attendees^2 3^ and adding to the burden on the healthcare system ^4^. Treatment of depression leads to improvement in functioning and reduction in healthcare expenses^5^. Longer duration of untreated illness negatively influences the course and outcome of depression^6–8^; however, more than 50% of potential cases of depression remain undetected in high income countries^2^. Among those whose depression is detected successfully and who are initiated on treatment, the majority have unstructured and inadequate access to treatment^9^. Due to the high prevalence and the significant level of disability attributable to depression, prioritizing the detection and management of depression and taking a public health approach is critical for multiple reasons^10^.

(i) Primary health care is the first entry point into the healthcare system and offers the best opportunity for illness and initiation of care^11 12^; (ii) patients generally prefer to be treated in primary care whenever they can; (iii) the primary care facility provides service users an accessible and relatively affordable opportunity for the receipt of healthcare for neglected health problems, including depression^11^; (iv) due to the predominant presentation with somatic symptoms, most people with depression visit primary care frequently^13^; (v) finally, the large treatment gap and the global effort to improve access to treatment for mental disorders^14 15^, makes PHC the most feasible and realistic platform for scaling up care in LMICs. The recognition of depression is, therefore, an important pre-requisite for reducing the treatment gap.

Given the overall importance of depression and its detection in PHC, this systematic review aimed to synthesize evidence about the detection of depression by PHC clinicians in LMICs, including factors that may facilitate or hinder detection. Additionally, the review aimed to evaluate the pooled prevalence of depression among the studies that assessed the detection of depression.

## METHODS

### Search strategy

The review protocol was registered in the PROSPERO database (CRD42016039704). Medline, PsycINFO, EMBASE and PubMed databases were searched since the inception of the respective databases until 3^rd^ Week of December 2019. AJOL and LILAC databases and manual search were also employed. The following terms were used to identify Depression: Depression OR depressive disorder OR Common mental disorder. The search terms used for detection were: Detection OR Detection rate OR Prevalence OR Screening OR Case finding OR Diagnosis OR Undiagnosed OR under-detection. For primary health care, we used Primary health care OR primary care OR Health centers. We used the World Bank definition and List of countries to identify LMICs. Terms for detection, depression, PHC and LMICs were combined with the Boolean term “AND” (Supplementary file 1).

### Outcomes of interest

The primary outcome was detection, defined as the proportion of the number of patients correctly diagnosed as having depression by primary care clinicians compared to a ‘gold’ standard diagnosis. The gold standard assessment included locally validated instrument or a confirmatory clinical diagnosis by a mental health expert. The secondary outcome was prevalence of depression among studies that reported detection. Factors associated with detection were also explored.

### Eligibility criteria

Eligible articles were assessed against the following inclusion criteria.

1. Diagnosis: both adults and adolescents aged 15 years and above with depression, including major depressive disorder, bipolar depression, masked depression, secondary depression, minor depression, and sub-threshold depression as determined by primary care clinicians irrespective of the offer of intervention.
2. Study Setting: LMICs at the time of the study, according to the World Bank classification^16^.
3. Type of study: Prospective studies, case-control studies, cross-sectional studies and clinical trials if aim to evaluate impact on detection.
4. Language: No language restriction.
5. Year of publication: primary studies published since the establishment of the respective databases until 3^rd^ Week of December 2019.

### Quality assessment

Risk of bias within the studies was assessed using the Effective Public Health Practice Project (EPHPP) quality assessment tool ^17^. The tool consists of eight quality assessment items: selection bias, allocation bias, control of confounders, blinding of outcome assessors, data collection methods, withdrawals and dropouts, intervention integrity, and analysis. The last three criteria were not included in overall rating because the review did not use intervention or analytical studies. Where follow-up studies were included, only the baseline data were used. Thus, five of the eight quality items were available for the overall rating. A global rating of ‘weak’, ‘moderate’, or ‘strong’ were made qualitatively. The global rating was rated ‘strong’ if no weak rating were given; moderate if only one weak rating and weak if two or more ‘weak’ ratings were made.

The quality of reporting was assessed using Strengthening the Reporting of Observational studies in Epidemiology (STROBE) checklist containing 22 items item ^18^ as a secondary assessment tool. We rated the 22 items by two of the authors (MD and RB) independently as per the STROBE guideline: “fully reported”, “partially reported” and “not reported”.

### Data extraction

Studies were first screened by two of the authors (MD and RB) independently, based on their titles and abstracts and any discrepancies were reconciled through discussions with a third author (AF). Excluded articles and reasons for exclusion were documented. Data were also extracted independently by the same two authors using a piloted data extraction form that included study country, study design, sample size, number of patients detected by clinicians, number of patients detected by the ‘gold’ standard tool, and the outcomes (detection, prevalence, associated factors).

### Statistical analysis

We conducted meta-analysis stratified by diagnostic approaches: diagnostic instrument (Beck Depression Inventory, Edinburgh Postnatal Depression Scale; PHQ with the two diagnostic thresholds; Structured Clinical Interview for DSM and the Composite International Diagnostic Interview based). Because of significant heterogeneity while pooling the outcome, a random effects meta-analysis was used. Pooled prevalence estimates were also obtained from the 18 studies stratified by the type of instrument used and the cutoff values of the instruments. Studies of special participant populations were excluded from the meta-analysis.

## RESULTS

A total of 4018 articles were identified. After removing 1795 duplicates, 2223 titles were reviewed, and 1895 articles were excluded. An additional 291 articles were excluded at the abstract review. A total of 37 articles were included in full article review and 10 publications with 18 country level reports were included in the final review (Figure 1).

**Figure 1:**
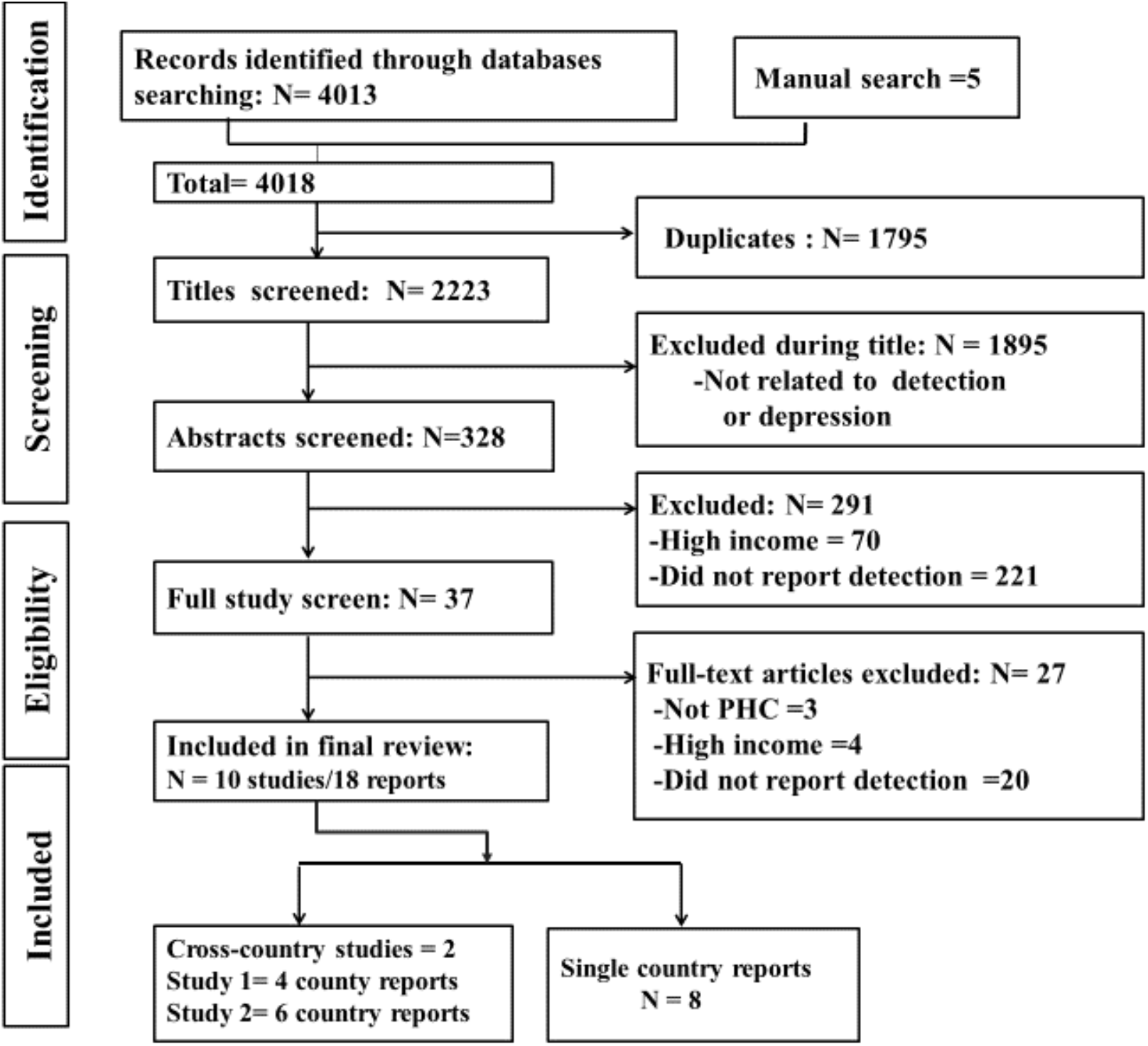
PRISMA Flow diagram of the study selection process.

### Characteristics of the included studies and case identification

The 10 studies or publications making the 18 individual country level reports came from two multi-country and eight single country studies (Table 1). The reports were published between 1995 and 2018 and had a total of 21,524 participants. Three reports each originated from India and Nigeria, two reports each from Ethiopia and Malawi, and one report each from Brazil, Chile, China, Nepal, Palestine, South Africa, Turkey and Uganda. All the studies except two were cross-sectional. The exceptions were one multi-country study, which was a follow up study and one cluster randomized controlled trial. For the later studies, only the baseline data were included.

**Table 1:**
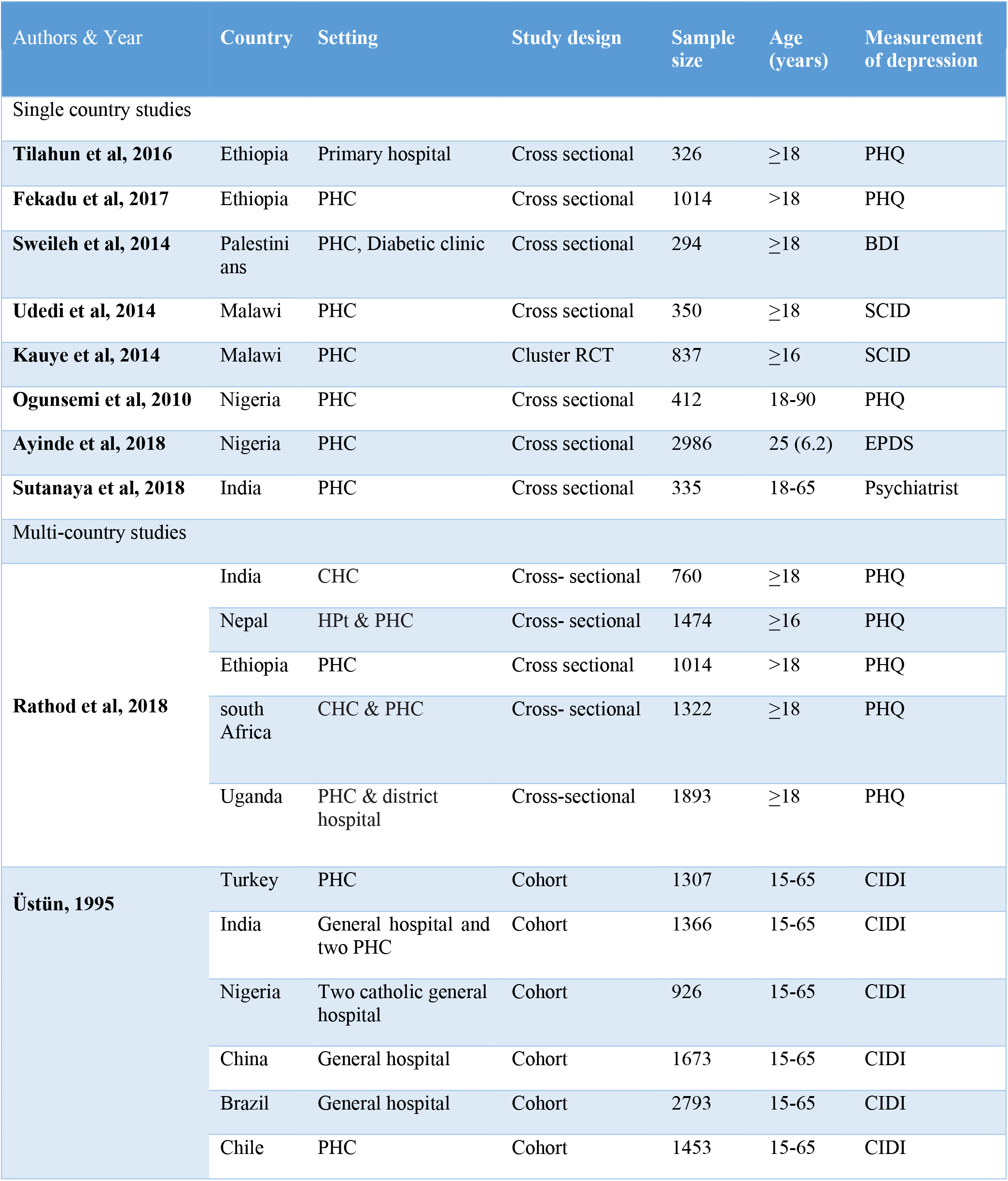
Key characteristics of the include studies.

| Authors & Year | Country | Setting | Study design | Sample size | Age (years) | Measurement of depression |
| --- | --- | --- | --- | --- | --- | --- |
| Single country studies |  |  |  |  |  |  |
| **Tilahun et al, 2016** | Ethiopia | Primary hospital | Cross sectional | 326 | ≥18 | PHQ |
| **Fekadu et al, 2017** | Ethiopia | PHC | Cross sectional | 1014 | >18 | PHQ |
| **Sweileh et al, 2014** | Palestinians | PHC, Diabetic clinic | Cross sectional | 294 | ≥18 | BDI |
| **Udedi et al, 2014** | Malawi | PHC | Cross sectional | 350 | ≥18 | SCID |
| **Kauye et al, 2014** | Malawi | PHC | Cluster RCT | 837 | ≥16 | SCID |
| **Ogunsemi et al, 2010** | Nigeria | PHC | Cross sectional | 412 | 18-90 | PHQ |
| **Ayinde et al, 2018** | Nigeria | PHC | Cross sectional | 2986 | 25 (6.2) | EPDS |
| **Sutanaya et al, 2018** | India | PHC | Cross sectional | 335 | 18-65 | Psychiatrist |
| Multi-country studies |  |  |  |  |  |  |
| **Rathod et al, 2018** | India | CHC | Cross- sectional | 760 | ≥18 | PHQ |
|  | Nepal | HPt & PHC | Cross- sectional | 1474 | ≥16 | PHQ |
|  | Ethiopia | PHC | Cross sectional | 1014 | >18 | PHQ |
|  | South Africa | CHC & PHC | Cross- sectional | 1322 | ≥18 | PHQ |
|  | Uganda | PHC & district hospital | Cross-sectional | 1893 | ≥18 | PHQ |
| **Üstün, 1995** | Turkey | PHC | Cohort | 1307 | 15-65 | CIDI |
|  | India | General hospital and two PHC | Cohort | 1366 | 15-65 | CIDI |
|  | Nigeria | Two catholic general hospital | Cohort | 926 | 15-65 | CIDI |
|  | China | General hospital | Cohort | 1673 | 15-65 | CIDI |
|  | Brazil | General hospital | Cohort | 2793 | 15-65 | CIDI |
|  | Chile | PHC | Cohort | 1453 | 15-65 | CIDI |

Regarding the setting, 12 reports were based in primary care clinics^19–23^, four studies in general hospitals where patients had direct access from the community^19 24^ and one primary care clinic for patients with diabetes ^25^ and one primary maternal care clinic^26^. One study recruited men only participants ^27^.

The professional background of the PHC clinicians varied across the studies depending on the health care structure and resources of each country and included health officers ^28^ with a medical training of four years^29^, medical officers, PHC doctors, nurses, health assistants and auxiliary health workers^19 20^. In the study reported from Brazil, the general health care centers were staffed by internists or residents^19^. Five studies did not report the background of the PHC clinicians ^21–25^. Regarding training, four studies (two cross-country studies and two additional individual studies) confirm that they did not provide additional mental health training or other intervention prior to the assessment^20 23 28 19^ while the remaining six studies do not provide such information ^21 22 24–27^.

Clinician diagnosis of depression was recorded in a clinician consultation or encounter form ^19 20 28^ or patients’ records were reviewed and diagnosis status extracted ^21 22 26 27^. Three reports did not indicate the methods they used to specify clinicians’ diagnosis ^23–25^.

Four studies, making nine reports, used two stage diagnostic screening to confirm the presence of depression^19 22 23^. In two of these studies ^22 23^, participants were first screened using the 20 item Self-Reporting Questionnaire (SRQ-20) and those who scored positive on the SRQ had a confirmatory diagnostic assessment using the Structured Clinical Interview for DSM-IV (SCID). The other study, a WHO multi-country study with six country reports, used the 12 item General Health Questionnaire (GHQ) for initial screening followed by confirmatory diagnosis with the primary care version of the Composite International Diagnostic Interview (CIDI) ^19^. Study participants in the second stage were all of the high scores, 35% of the medium scores and 10% of the low scorers. The fourth study used PHQ-9 followed by a psychiatrist assessment for confirmation of depression ^27^.

The remaining studies relied on a single assessment--the 9-item Patient Health Questionnaire (PHQ-9) ^20 21 24^ or the Beck Depression Inventory (BDI-II) ^25^ and the Edinburgh Postnatal Depression Scale^26^--for the diagnosis of probable depression. For the PHQ, varied cutoff scores were used as threshold for detection: a cutoff of threshold score of 10 ^20^ and score of 5 ^21 24^ and both scores of 5 and 10 ^28^. One study also computed DSM-V Major Depressive Disorder (MDD) from the PHQ ^28^.

Fifteen out of the 18 reports noted the order of assessment in relation to primary care clinician. In the reports that relied on a two stage diagnosis (n=9), the initial screening instrument was administered before clinician assessment ^22 23^. For the reports that used a single stage screening (n=6): two studies offered screening before the participants had clinician assessment but, the clinicians were blind to the results of the screening ^20^; in the other four reports screening happened after assessment by clinician ^20 21^.

### Quality of included studies

According to the EPHPP quality assessment, four studies were assessed to be of strong quality, five of moderate quality and only one study of weak quality (Supplementary file 2). The overall quality of reporting of the studies was also moderate to high as per the STROBE checklist (Supplementary file 3).

### Detection by primary care clinicians

In all studies, detection was defined as the proportion of patients diagnosed as having depression by primary care clinicians among participants who screened positive using standard assessment scale or instrument. The detection was 0% in five of the 18 reports ^20 22–25^ and under 10% in another four reports ^20 28^.

The pooled detection level from the two reports that used PHQ-9 with a cutoff score of 5 as a gold standard was 3.9% (95% CI = 2.3%, 5.5%). For the four reports that used PHQ-9 at a cutoff score of 10 as a ‘gold standard’, the pooled detection level was 7.0 % (95% CI = 3.9%, 10.2%), with no significant heterogeneity (I^2^ = 43.5%, P= 0.15). For the WHO study, the pooled estimate of detection of depression from the six LMICs was 43.5 % (95% CI: 25.7%, 61.2%) (Figure 2 and 3) and that of dysthymia was 63.5% (95% CI: 47.0%, 80.1%).

**Figure 2:**
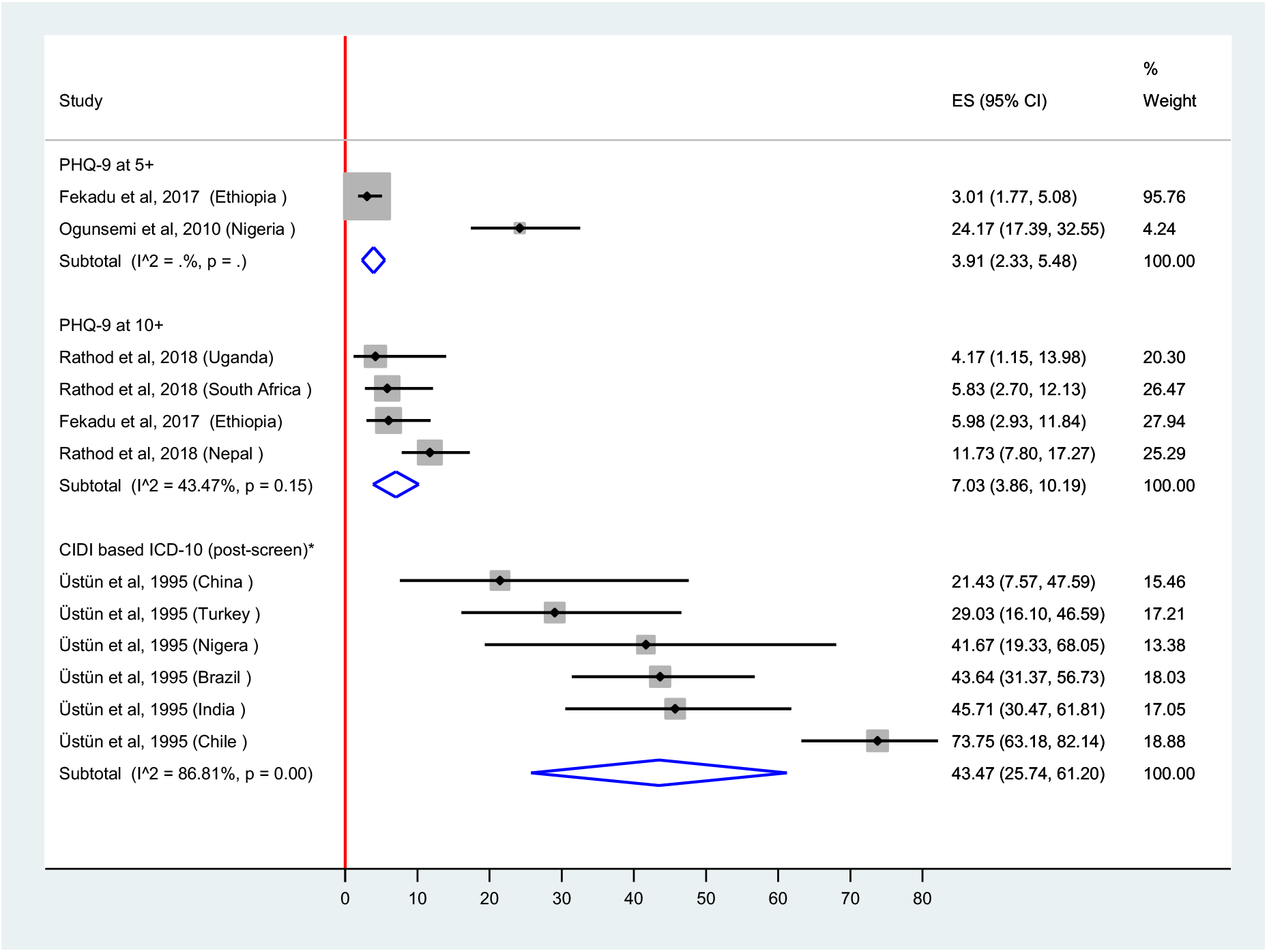
Pooled estimate of detection stratified by diagnostic approaches using a random effects meta-analysis.

**Figure 3.**
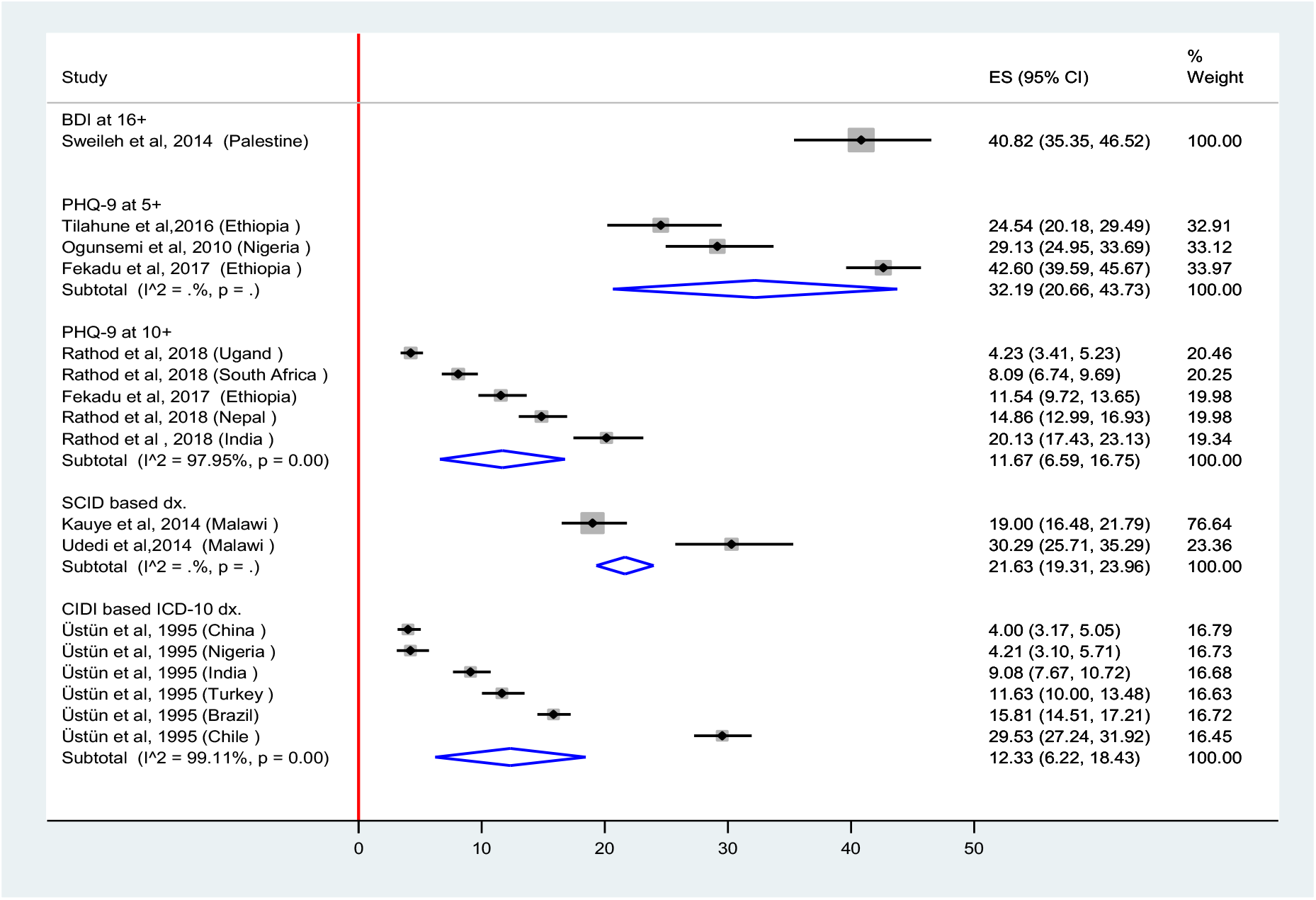
Pooled prevalence of depression stratified by diagnostic approaches using random effects model.

The study among men adult clinic attendees reported detection of 69.0% with just two physician assessors^27^. The study carried out among pregnant women who were joining ANC follow up reported a detection of 1.4% (n=3/218)^26^.

Only one study evaluated factors associated with detection and reported severity of depression and suicidality to be associated with detection. Additionally, although not significant, women and those with higher educational attainment were more likely to be diagnosed with depression ^28^ (Figure 2).

### Prevalence of depression in the included studies

The pooled prevalence of depression (Figure 3) in reports that used PHQ-9 at a cutoff score of 10 was 11.7 % (95% CI: 6.6%, 16.8%) and it was 32.2% (95% CI: 20.7 %, 43.7%) among studies that used a score of 5 as a cutoff. One study reported prevalence of 40.8 % (95% CI: 35.4%, 46.5%) using BDI at a cutoff score of 16. The pooled prevalence of depression was 21.6 % using SCID based DSM criteria. The pooled prevalence of major depressive disorder from three studies reporting on the disorder was 9.3% (95% CI= 2.4%, 16.2%). In the WHO multi-country study that used the CIDI, the pooled prevalence of depressive disorder was 12.3% (6.2%, 18.4%) and that of dysthymia was 3.3% (1.52%, 4.55%).

It was not possible to include estimates from the three studies conducted in special participant groups in the pooled estimate. The reported prevalence figures were as follows: 40.8 % (95% CI: 35.4%, 46.5%) among people with Diabetics ^25^, 7.3% among pregnant women who were on ANC follow-up ^26^ and 22% among male only participants^27^.

## DISCUSSION

This is the first systematic review and meta-analysis of studies reporting on the detection of depression by primary care clinicians in LMICs. Overall, we identified very limited number of studies reporting on the subject. Where studies existed, the level of detection was low, with 0.0% detection in five reports ^20 22–25^ and another four reporting levels under 10%^20 28^. Detection was better in studies that used enriched sample where participants were first screened for depression before being evaluated by primary care clinicians. Even in these studies, the pooled detection estimate was under 45%, with individual detection reports ranging from about 20% to 70%.

This level of detection compares unfavourably against studies from high income countries^2^, where average detection level is around 50%. The low detection cannot be explained by a low prevalence. The pooled prevalence of possible depression in this review suggests that up to a third of primary care attendees may have depression, which is consistent with the broader literature ^19^. In fact, a recent systematic review reports higher prevalence of depression and depressive symptoms among outpatients in developing countries than outpatients from developed countries^30^. The low detection is a reflection of the broader neglect of people with depression in LMICs. For example, a recent global report on adequacy of treatment for depression showed that only about 4% of people with depression receive minimally adequate treatment in LMICs compared with 20% in non-LMICs^31^. In the context of the need to scale up mental health services in LMICs and reduce the treatment gap ^32^, this low detection rate should be of major concern. Within this scale up initiative, people with depression stand to benefit the most given the high prevalence of depression in PHC and its overall treatment responsiveness. In essence, the low detection means that most people with mental health problems who would benefit from treatment in primary care are not benefiting^33^.

Generally, depression is under-detected and undertreated in primary care globally, even in high income countries ^2 34^. However, health system factors, such as longer engagement in care, offer a better prospect for detection in high income countries ^35^. Improving detection should be one of the research priorities, there is no robust evidence in LMICs on how best to improve detection. The only study indicating benefit of an intervention, a training-based approach conducted in sub-Saharan Africa^23^, has not been replicated. Moreover, a similar intervention in Kenya did not improve the rate of detection ^36^. Five of the studies included in this review were part of a lager study that aimed at developing the best evidence on integrating mental healthcare, including depression care, into primary care using the latest WHO guideline^32^. In this regard, a novel and complex approach is required to enhance detection and engagement in care.

Only one of the included studies reported on predictors of detection, which noted severity of symptoms and suicidality as significant predictors^28^. Overall, these are consistent findings with what is known about the detection of depression ^37^. Thus, the main challenge remains improving the threshold for detection, although studies on determinants of detection may be informative.

However, the review has several limitations. First, there were only few studies available for this review. Second, there was high heterogeneity. Third, screening tools were used as gold standard diagnosis. This is likely to inflate the prevalence of depression and underestimate the detection rate. We also focused on depression and not broader psychiatric morbidity or common mental disorders, which may be the overriding presentation in PHC. The limitation of the binary approach in the research and discourse around depression and mental disorders is well recognized^38^. In relation to the individual studies, some of the studies have low sample size^24 25^, and this may be too low to provide reliable estimate of detection.

## CONCLUSION

To our knowledge, this review is the first attempt to bring together all relevant studies of the detection of depression in primary care in LMICs. The review shows the dearth of studies on detection of depression in primary care and also highlights the challenges of improving detection. Improving detection of depression should be an important next step in scaling up services in LMICs. Depression is considered rightly a ‘priority development challenge’^38^ and the low rate of depression is an important barrier to addressing this development challenge. Interventions that improve detection rate by primary care clinicians in LMICs and studies on factors associated with detection are warranted.

## Data Availability

Data sharing not applicable as no datasets generated and/or analysed for this study

## Acknowledgments

GT is supported by the National Institute for Health Research (NIHR) Collaboration for Leadership in Applied Health Research and Care South London at King’s College London NHS Foundation Trust. The views expressed are those of the author(s) and not necessarily those of the NHS, the NIHR or the Department of Health. GT also receives support from the National Institute of Mental Health of the National Institutes of Health under award number R01MH100470 (Cobalt study). GT is also supported by the UK Medical Research Council in relation the Emilia (MR/S001255/1) and Indigo Partnership (MR/R023697/1) awards.

